# Exome sequencing identifies novel susceptibility genes and defines the contribution of coding variants to breast cancer risk

**DOI:** 10.1101/2022.06.17.22276537

**Authors:** Naomi Wilcox, Martine Dumont, Anna González-Neira, Sara Carvalho, Charles Joly Beauparlant, Marco Crotti, Craig Luccarini, Penny Soucy, Stéphane Dubois, Rocio Nuñez-Torres, Guillermo Pita, M. Rosario Alonso, Nuria Álvarez, Caroline Baynes, Heiko Becher, Sabine Behrens, Manjeet K. Bolla, Jose E. Castelao, Jenny Chang-Claude, Sten Cornelissen, Joe Dennis, Thilo Dörk, Christoph Engel, Manuela Gago-Dominguez, Pascal Guénel, Andreas Hadjisavvas, Eric Hahnen, Mikael Hartman, Belén Herráez, SGBCC Investigators, Audrey Jung, Renske Keeman, Marion Kiechle, Jingmei Li, Maria A. Loizidou, Michael Lush, Kyriaki Michailidou, Mihalis I. Panayiotidis, Xueling Sim, Soo Hwang Teo, Jonathan P. Tyrer, Lizet E. van der Kolk, Cecilia Wahlström, Qin Wang, Javier Benitez, Marjanka K. Schmidt, Rita K. Schmutzler, Paul D.P. Pharoah, Arnaud Droit, Alison M. Dunning, Anders Kvist, Peter Devilee, Douglas F. Easton, Jacques Simard

**Author notes:** These authors jointly supervised this work (Douglas F. Easton, Jacques Simard).

## Abstract

Linkage and candidate gene studies have identified several breast cancer susceptibility genes, but the overall contribution of coding variation to breast cancer is unclear. To evaluate the role of rare coding variants more comprehensively, we performed a meta-analysis across three large whole-exome sequencing datasets, containing 16,498 cases and 182,142 controls. Burden tests were performed for protein-truncating and rare missense variants in 16,562 and 18,681 genes respectively. Associations between protein-truncating variants and breast cancer were identified for 7 genes at exome-wide significance (*P*<2.5×10^-6^): the five known susceptibility genes *BRCA1, BRCA2, CHEK2, PALB2* and *ATM*, together with novel associations for *ATRIP* and *MAP3K1*. Predicted deleterious rare missense or protein-truncating variants were additionally associated at *P*<2.5×10^-6^ for *SAMHD1*. The overall contribution of coding variants in genes beyond the previously known genes is estimated to be small.

## Main Text

Breast cancer is the leading cause of cancer-related mortality for women worldwide. Genetic susceptibility to breast cancer is known to be conferred by common variants, identified through genome-wide association studies (GWAS), together with rarer coding variants conferring higher disease risks. The latter, identified through genetic linkage or targeted sequencing studies, include protein-truncating variants (PTVs) and/or some rare missense variants in *ATM, BARD1, BRCA1, BRCA2, CHEK2, RAD51C, RAD51D, PALB2* and *TP53*^*1*^. However, these variants together explain less than half the familial relative risk (FRR) of breast cancer^2^. The contribution of rare coding variants in other genes remains largely unknown.

Here, we used data from three large whole-exome sequencing (WES) studies, primarily of European ancestry, to assess the role of rare variants in all coding genes: the BRIDGES (Breast Cancer Risk after Diagnostic Gene Sequencing) dataset that included samples from eight studies in the Breast Cancer Association Consortium (BCAC), the PERSPECTIVE (Personalised Risk assessment for prevention and early detection of breast cancer: integration and implementation) dataset that included three BCAC studies (Supplementary Table 1), and UK Biobank. After quality control (see methods), these datasets comprised 16,498 cases and 182,142 controls (Supplementary Table 2).

We considered two main categories of variants: protein-truncating variants (PTVs) and rare missense variants (minor allele frequency <0.001). Single variant association tests are generally underpowered for rare variants; however, burden tests, in which variants are collapsed together, can be more powerful if the associated variants have similar effect sizes^3^. To further improve power, we incorporated data on family history of breast cancer (see methods)^4^. Association tests were conducted for all genes in which there was at least one carrier of a variant (16,562 genes for PTVs and 18,681 genes for rare missense variants).

In the combined analysis of PTVs, 18 genes were associated at P<0.001 (Supplementary Table 3, Figures 1-2). Of these, 7 met exome-wide significance (P<2.5×10^-6^), of which 5 are known breast cancer risk genes - *BRCA1, BRCA2, CHEK2, PALB2 and ATM*. Novel associations were identified for PTVs in *MAP3K1* (P=6.1×10^-8^) and *ATRIP* (P=1.8×10^-6^). Of the other previously identified breast cancer susceptibility genes, nominally significant associations were observed for *BARD1, CDH1* and *RAD51D* (Supplementary Table 4). There was no evidence for an excess of significant associations after allowing for the 7 exome-wide significant genes (Figure 2). However, 17 of the 18 associations at P<0.001 correspond to an increased risk, compared with ∼9.5 that would be expected by chance. This imbalance suggests some of the other associations may be genuine.

**Figure 1.**
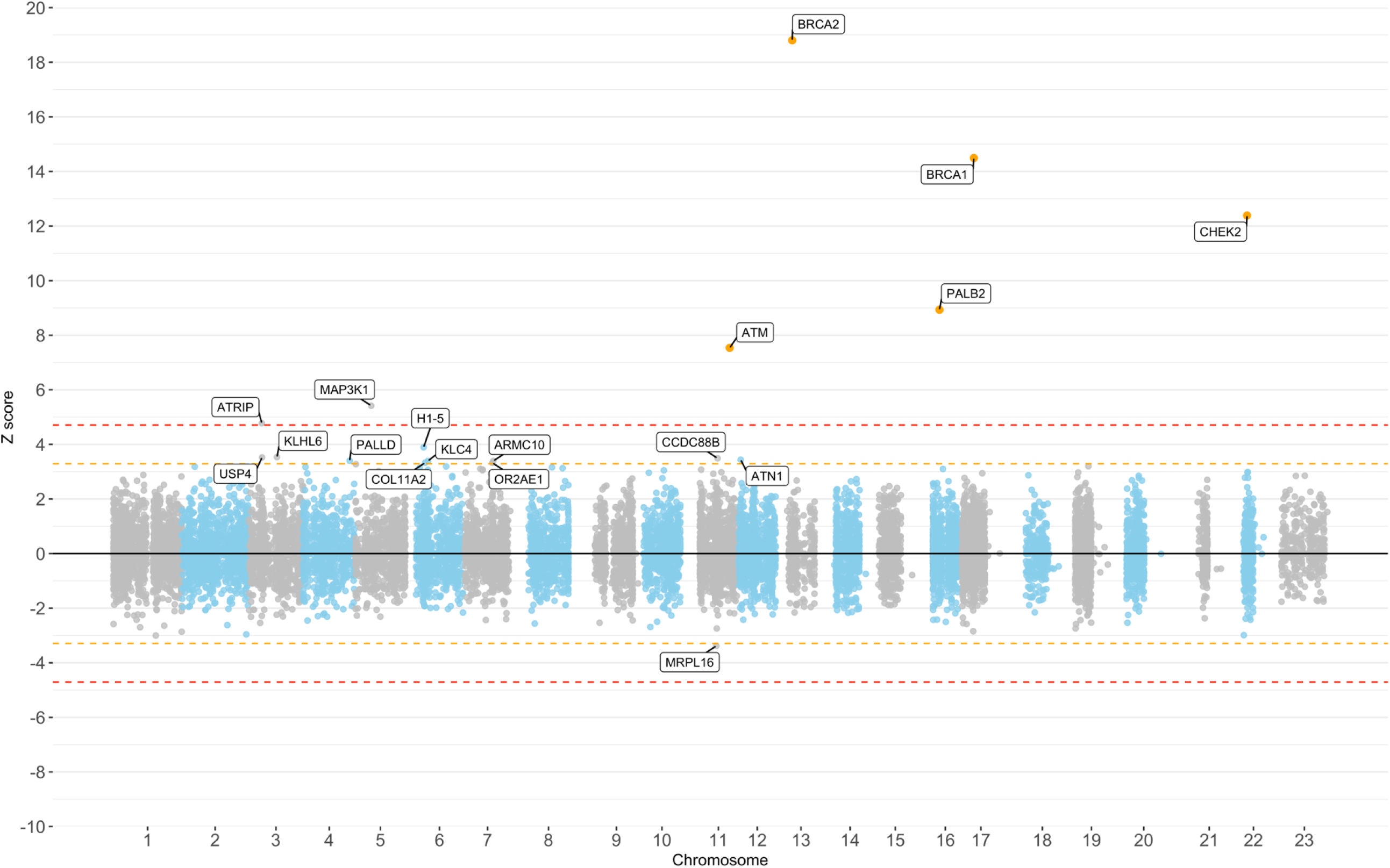
Manhattan Plot of Z-scores from the meta-analysis assessing the association between PTV carriers within genes and breast cancer risk. The orange line corresponds to Z=±3.29, p=0.001. Red line corresponds to Z=±4.71, p=2.5×10^-6^.

**Figure 2.**
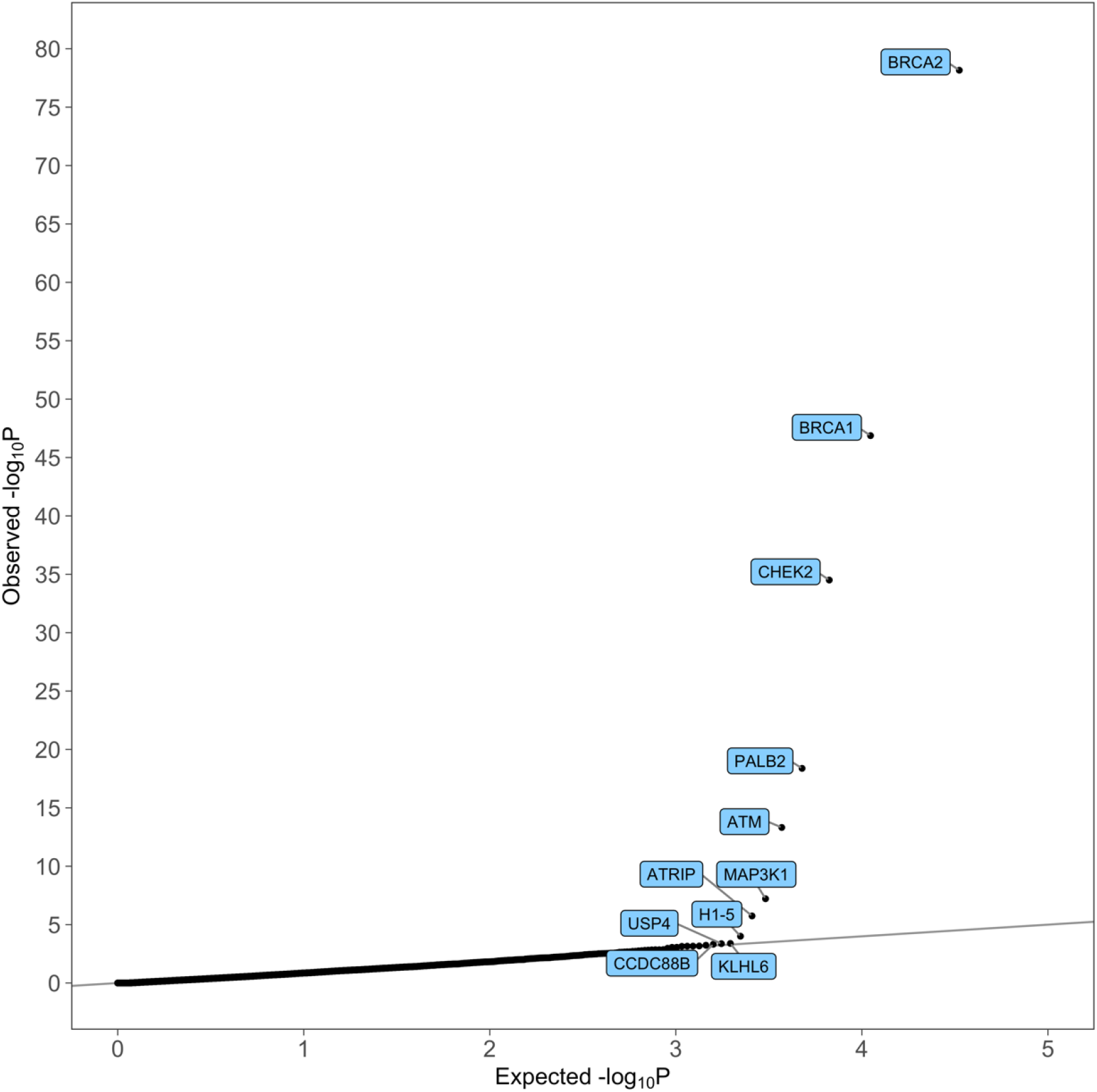
Quantile-Quantile Plot of P-values from the meta-analysis assessing the association between PTV carriers and breast cancer risk. All highlighted genes with p<0.0005 correspond to an increased risk of breast cancer.

Within the BCAC dataset, we evaluated the association with subtypes of breast cancer; ER+ or ER-, PR+ or PR-, and triple-negative disease. We compared different subtypes of cases to controls and performed analysis within cases only (Supplementary Table 5). The expected associations for known genes were observed, notably the higher OR for ER-negative and triple-negative disease for *BRCA1* and higher OR for ER-positive disease for *CHEK2*, but no additional genes were associated with subtype-specific disease at exome-wide significance.

For the rare missense variant meta-analysis, 15 genes had a P-value <0.001, 12 of which corresponded to an increased risk of breast cancer (Supplementary Table 6, Supplementary Figures 1-2) compared to 7.5 expected by chance. Only *CHEK2* met exome-wide significance (P=1.7×10^-11^). We next considered missense variants predicted deleterious by CADD (CADD score ≥20) combined with PTVs. In this analysis, 24 genes had a P-value < 0.001, 17 of which corresponded to an increased risk of breast cancer (Supplementary Table 7); Supplementary Figures 3-4). Six genes met exome-wide significance: *CHEK2* (P=2.1×10^-42^), *BRCA2* (P=6.3×10^-18^), *PALB2* (P=1.1×10^-9^), *BRCA1* (P=1.5×10^-9^), *ATM* (P=3.4×10^-7^), and *SAMHD1* (P=5.8×10^-7^).

Notably, all three novel genes achieving exome-wide significance, *MAP3K1, ATRIP* and *SAMHD1* have prior evidence of involvement in tumorigenesis or DNA repair. *MAP3K1* is a stress-induced serine/threonine kinase that activates the ERK and JNK kinase pathways by phosphorylation of MAP2K1 and MAP2K4^5,6.^ Inactivating variants in *MAP3K1* are one of the commonest somatic driver events in breast tumors^7,8^. *MAP3K1* is also a well-established GWAS locus^9^; at least 3 independent signals have been identified mapping to regulatory regions with *MAP3K1* expression as the likely target^10,11^. To evaluate whether the *MAP3K1* PTV association we observed was driven by the GWAS associations, or vice-versa, we fitted logistic regression models to UK Biobank data in which the PTV burden variable and the lead GWAS SNPs (SNP_1_: rs62355902, SNP_2_: rs984113 and SNP_3_: rs112497245) were considered jointly (Supplementary Table 8). In the model with all variables, the OR associated with carrying a PTV (OR = 8.54 (2.96, 24.66)) was similar to the unadjusted OR. Similarly, the ORs for each of the SNPs were similar to the ORs without adjustment for PTVs. This suggests that the PTV burden and GWAS associations are independent and reflect the distinct effects of inactivating coding alterations and regulatory variants.

*ATRIP* (ATR interacting protein) codes for a DNA damage response protein which forms a complex with ATR. ATR-ATRIP is involved in the process that activates checkpoint signalling when single-stranded DNA is detected following the processing of DNA double-stranded breaks or stalled replication forks^12,13.^ *SAMHD1* promotes the degradation of nascent DNA at stalled replication forks, limiting the release of single-stranded DNA^14^. *SAMHD1* also encodes dNTPase that protects cells from viral infections^15^ and is frequently mutated in multiple tumor types, including breast cancer.

Pathology information was available in the BCAC dataset for 9 carriers of *MAP3K1* PTVs, 14 carriers of *ATRIP* PTVs and 46 carriers of *SAMHD1 PTVs or* predicted deleterious rare missense variants. These data suggest a higher proportion of low-grade, low-stage ductal breast cancer for *MAP3K1* carriers, but high-grade, low-stage ductal breast cancer for *ATRIP* carriers (Supplementary Table 9). However, none of the associations with tumor characteristics were statistically significant.

To evaluate the overall contribution of PTVs to the Familial Relative Risk (FRR), we fitted models to the effect size using an empirical Bayes approach. Under the assumption of an exponentially distributed effect size, the estimated proportion of risk genes was 0.0066 with a median OR of 1.38. Under this model, an estimated 18.07% of the FRR would be explained, of which 16.01% would be due to the five genes *BRCA1, BRCA2, ATM, CHEK2* and *PALB2* and 2.06% due to all other genes combined (*MAP3K1* – 0.44%, *ATRIP* – 0.13%) (Supplementary Table 10). Only the seven genes reaching exome-wide significance for PTVs had a posterior probability of association >0.90 (with all other posterior probabilities <0.4).

These results demonstrate that large exome sequencing studies, combined with efficient burden analyses, can identify additional breast cancer susceptibility genes. The excess of positive associations at P<0.001 indicates that further genes should be identifiable through large datasets: the heritability analyses suggest the number of associated genes might be of the order of 130. Further replication in larger datasets will also be necessary to provide more precise estimates for variants in the novel genes *ATRIP, MAP3K1* and *SAMHD1*, to define the set of variants in these genes associated with breast cancer and the clinic-pathological characteristics of tumors in variant carriers. The heritability analyses suggest that most of the contribution of PTVs is mediated through the five genes *BRCA1, BRCA2, ATM, CHEK2* and *PALB2*, commonly tested for in clinical cancer genetics^16^. While subsets of missense variants may also make important contributions (exemplified by *SAMHD1*), these results suggest that the majority of the “missing” heritability is likely to be found in the non-coding genome.

## Data Availability

Data from UK Biobank are available through application to UK Biobank. Data from the Breast Cancer Association Consortium (BCAC) used in the present study are available upon reasonable request through the BCAC Data Access Co-ordinating Committee.

## Acknowledgements

Sequencing and analysis for this project were funded by the European Union’s Horizon 2020 Research and Innovation Programme (BRIDGES: grant number 634935), the PERSPECTIVE I&I project (funded by the Government of Canada through Genome Canada and the Canadian Institutes of Health Research, the Ministère de l’Économie et de l’Innovation du Québec through Genome Québec, the Quebec Breast Cancer Foundation, Agilent Technologies Canada Inc and Illumina Canada Ulc) and the Wellcome Trust [grant no: v203477/Z/16/Z]. BCAC is funded by the European Union Horizon 2020 Research and Innovation Programme (grant numbers 634935 and 633784 for BRIDGES and B-CAST respectively), the PERSPECTIVE I&I project and via the Confluence project which is funded with intramural funds from the National Cancer Institute Intramural Research Program, National Institutes of Health. The funders had no role in the study design, data collection, data analysis, data interpretation or writing of the report. This research has been conducted using the UK Biobank Resource under Application Number 28126. BCAC study-specific funding is given in the Supplementary Note.

## Online Methods

### UK biobank

The UK Biobank is a population-based prospective cohort study of more than 500,000 samples. More detailed information on the UK Biobank is given elsewhere^17,18^. WES data for 200,000 samples were released in October 2020^19^. QC metrics were applied to Variant Call Format files^19^. At the genotype level, SNPs were excluded with sequencing depth <7 or heterozygous allele balance <0.15 or >0.85. Indels were excluded with sequencing depth <10 or allele balance <0.2 or >0.8. On males’ X chromosomes, depth filters were reduced to 5 for SNPs and 7 for indels. Samples with missing calls for >15% and variants with missing calls for >15% of samples were excluded. Variants with Hardy Weinberg Equilibrium p-value 10^-15^ were also removed.

Samples with disagreement between genetically determined and self-reported sex, sex aneuploidy, or excess relatives in the dataset were excluded. Excess relatives were identified by considering pairs of individuals with kinship>0.17. If one individual in a pair was a case and one was a control then the case was preferentially selected; otherwise, one individual was selected at random. Genetic ancestry was estimated using genetic principal components and the Gilbert-Johnson-Keerthi distance algorithm^20^. If genetic principal components were not available, self-reported ethnicity was used. Samples of ancestry other than European were excluded. The final dataset for analysis included 181,992 samples with 100,068 Females. Cases were defined by having invasive breast cancer (ICD-10 C50) or carcinoma-in-situ (D05), as determined by linkage to national cancer registration (NCRAS), or self-reported breast cancer. Both prevalent and incident cases were included. Only breast cancers which were an individual’s first or second diagnosed cancer were included as cases. By this definition, 8,043 female and 45 male cases were included.

### The Breast Cancer Association Consortium datasets

The BRIDGES and PERSPECTIVE samples were from studies in the BCAC (BRIDGES: 8 studies, PERSPECTIVE: 3 studies; Supplementary Table 1). Most samples were previously included in a targeted panel sequencing project^1^. Phenotype data were based on the BCAC database v14. Samples were oversampled for early-onset (age of diagnosis below 50 years) or family history of breast cancer. Cases were preferentially selected to have information on tumor pathology. Samples with previously identified pathogenic mutations in *BRCA1, BRCA2* or *PALB2* were excluded.

For BRIDGES, library preparation was conducted in the 3 laboratories using the Nextera DNA Exome kit (Illumina) for tagmentation, barcoding and amplification steps. Subsequently, 500ng of DNA per sample were pooled in 12-plex and concentrated using a vacuum system. Afterwards, hybridization capture reagents for DNA libraries were used for overnight hybridization with the xGen Exome Research panel (Integrated DNA technologies, IDT), capture and amplification. Barcoded pooled libraries of 96 samples were sequenced on each lane of a NovaSeq 6000 S4 flowcell (Illumina) using NovaSeq Xp 4-Lane Kit (2×100bp).

For PERSPECTIVE, library preparation was conducted using Agilent SureSelect Human all exon V7 (48.2Mb). Barcoded libraries of 88 samples were sequenced on a NovaSeq 6000 S4 flowcell (Illumina) using NovaSeq Xp4-lane Kit (2×100bp).

The same pipeline for variant calling was applied to both the BRIDGES and PERSPECTIVE data and followed the GATK (Genome Analysis Toolkit) best practices. Briefly, raw sequence data (FASTQ format) were pre-processed to produce BAM files. This involved alignment to the reference genome, identification and removal of duplicate read pairs from the same DNA fragments, and base recalibration. The base recalibration included the generation of a base quality score recalibration table, later applied to the read bases to adjust their quality scores and increase the accuracy of the variant calling algorithms. An intermediate and informal Quality Control was performed for a sanity check, including coverage and alignment mapping metrics. Variants were then called using Haplotype Caller for the whole exome. This was later split into chromosomes for the analysis due to file size constraints. Variants were further filtered using Variant Quality Score Recalibration (VQSR), a proprietary algorithm from GATK that applied new calibration scores independently at SNP and indel variant levels.

From the final dataset, samples and variants were excluded based on coverage, allelic balance, and Hardy-Weinberg equilibrium, using the same filtering as for UK Biobank. We also excluded samples where the genotypes were inconsistent with previous array genotyping or targeted sequencing data^1,2.^

The BRIDGES study sequenced 6,912 samples, of which 3,461 cases and 3,200 controls remained in the final dataset after QC. The PERSPECTIVE study sequenced 10,523 samples, of which 4,777 cases and 5,210 remained in the final dataset.

### Data preparation

For both the UK Biobank and BCAC datasets, Ensembl Variant Effect Predictor (VEP) was used to annotate variants^21^. Annotations included the 1000 genomes phase 3 allele frequency, sequence ontology variant consequences and exon/intron number. For each gene, the MANE Select^22^ transcript was used if it was available for that gene, or the RefSeq Select transcript^23^. Annotation files were used to identify PTVs and rare (allele frequency <0.001 in both the 1000 genomes dataset and the current dataset) missense variants. PTVs in the last exon of each gene were excluded as these are generally predicted to escape Nonsense-Mediated mRNA Decay (NMD). VEP was also used to annotate missense variants by Combined Annotation Dependent Depletion score (CADD)^24^. Here CADD ≥20 was used to define variants predicted to be deleterious.

### Burden Test analysis

Association analyses were carried out for each gene separately for PTVs, rare missense variants, and predicted deleterious rare missense variants (defined by CADD score ≥20) and PTVs combined. The main association analyses were burden tests in which genotypes were collapsed to a 0/1 variable based on whether samples carried a variant of the given class. That is, 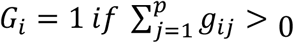 *and* 0 *if* 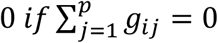 where *g*_*ij*_ = 0, 1, 2 is the number of minor alleles observed for sample i at variant j, and p is the number of variants in the gene (thus, heterozygous and homozygous carriers were combined).

Different statistical tests were considered by simulation, and the best method was chosen for each dataset. For the BCAC datasets, we used an exact conditional test^25,26,^ stratified by country and library preparation method (BRIDGES vs Perspective). Ethnicity was not adjusted for as within each country ethnicity was constant. This method had greater power than the Mantel-Haenszel test^27^ and Wald test from logistic regression^28^, and the type-1 error rate was closest to the specified significance level. The exact conditional test was also used for case-control analyses for subtypes and case-only analysis when comparing subtypes e.g., oestrogen receptor status.

Two genes with very common PTVs, *NUDT11* and *ZNF598*, were excluded because missing genotypes (which were treated as non-carriers) led to spurious associations even though the variants passed QC filtering. *AFF1* was also removed as the PTV frequency was high in PERSPECTIVE but rare in BRIDGES and UKB. This was likely due to a single PTV artefact within the PERSPECTIVE dataset.

For UK Biobank, we used logistic regression analysis but also incorporated family history as a surrogate for disease status. This markedly improves power since susceptibility variants will also be associated with family history; in particular, it allows information on males in the cohort with a family history of female breast cancer to be utilised. To do this, we treated genotype (0/1) as the dependent variable and family history weighted disease status as the covariate; the latter is defined as d+1/2f, where d=0,1 was the disease status of the genotyped individual and f=0 or 1 according to whether the individual reported a positive first-degree family history. The rationale for this weighting is that, for small effect sizes, the log(odds ratio) associated with a positive first-degree relative is approximately ½ that associated with the disease. All analyses adjusted for the first 10 principal components. For genes on chromosome X, only females were used in the analysis.

To combine the results from the BCAC and UK Biobank datasets in a meta-analysis, the association tests for each gene were converted to Z-scores. The combined Z-score was defined as: 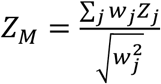. Here, *Z*_*M*_ is the combined z-score, *Z*_*j*_ is the z-score for study j and *w*_*j*_ is the weight associated with study j.

A standard meta-analysis would define the weights *w*_*j*_ using inverse variance or effective sample sizes. However, the effect sizes from the BCAC and UK Biobank may not be comparable, since the BCAC studies oversampled for family history and early age at onset, which may have increased the estimated effect. Furthermore, the UK biobank analysis incorporated family history, which changes the effective sample size as well as the effect estimate. Therefore, we defined weights by using the associations in the known risk gene *CHEK2* as a standard: we rationalised that the *CHEK2* PTVs provided the best standard as the association is well established and the odds ratio is highly reproducible^1,29^,30. Moreover, the odds ratio (∼2-2.5) was representative of the size of effects we hoped to detect for other genes. Thus, we defined 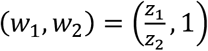, where 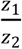 is the ratio of z-scores for *CHEK2* for the BCAC dataset and UK Biobank. A similar approach was applied for the meta-analysis of the other variant categories. *Z*_*M*_-scores were plotted in Manhattan plots and associated P-values plotted in quantile-quantile plots. The lambda statistic for inflation in the test statistics (based on the median chi-squared statistic) was 0.638 for UK Biobank, 0.549 for BCAC and 0.571 for the meta-analysis, indicating that the tests were somewhat conservative on average.

To investigate the joint effect of PTVs in *MAP3K1* and common susceptibility variants in the region identified through GWAS, we accessed imputed genotype data from UK Biobank for the lead SNPs as identified through previous fine-mapping analyses^10,11^ We fitted logistic regression models including covariates for PTVs and the lead SNPs and compared the fit of the model, and effect sizes, with the model in which the PTVs or the lead SNPs were excluded.

Data on clinicopathological characteristics of cases in the BCAC dataset was also accessed and the proportion of individuals with specific pathologic features e.g., stage and grade were compared between carriers of variants in a specific gene, e.g., *MAP3K1* PTV carriers, and the overall dataset.

### Contribution of PTVs to the FRR

We estimated the overall contribution of PTVs to the familial relative risk (FRR) of breast cancer using an empirical Bayesian approach. Given the aggregate frequency *p*_*j*_ of PTVs in a gene is rare, and all PTVs confer the same relative risk 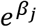, the FRR due to one gene, given *p*_*j*_ and 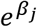, is:

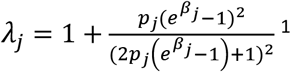

Under the additional assumption that the risks conferred by variants in different genes are additive, the total contribution over J genes is given by:

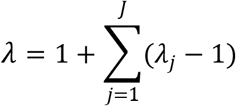

We assumed a prior distribution for effect sizes (log-odds ratios) in which a proportion *α* of genes are risk associated, and the estimated log-odds ratios *β* for associated genes have an exponential distribution with parameter *η*; this distribution was chosen since the distribution of effect sizes is likely to be skewed, with only a small number of genes have a large effect size and most undiscovered genes having smaller effect sizes. An approximate likelihood of the observed carrier count data, by gene, was derived, summed over all genes and maximised numerically to estimate *α* and *η*, and hence posterior effect size distributions given the data. The total contribution to the FRR was estimated by integrating the FRR estimates given *β*_*j*_ over the posterior distribution. Further details are given in Supplementary Methods.

## Notes

### Competing Interest Statement

The authors have declared no competing interest.

### Funding Statement

The work was funded by grants from:
Genome Canada
Genome Quebec
Canadian Institutes of Health Research
European Union Horizon 2020 Research and Innovation Programme
Wellcome Trust

### Author Declarations

A list of ethics committees is provided as a separate supplementary table.

