## Supplementary Figures 1-4 for "Exome sequencing identifies novel susceptibility genes and defines the contribution of coding variants to breast cancer risk"

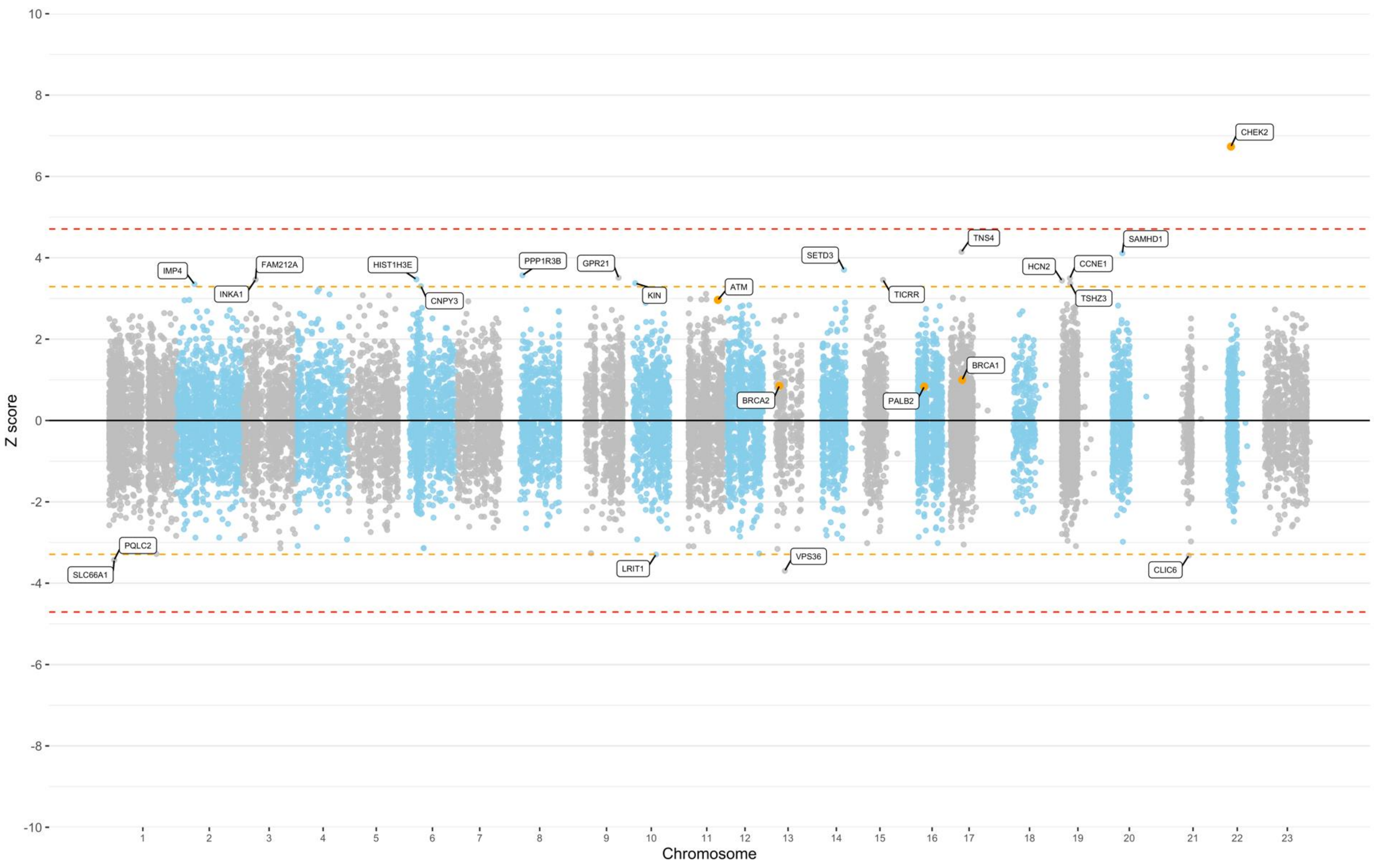


**Supplementary Figure 1 | Manhattan plot of Z-scores from the meta-analysis assessing the association between rare missense variant carriers within genes and breast cancer risk.** Orange line corresponds to Z=±3.29, p=0.001 Red line corresponds to Z=±4.71, p=2.5x10-6

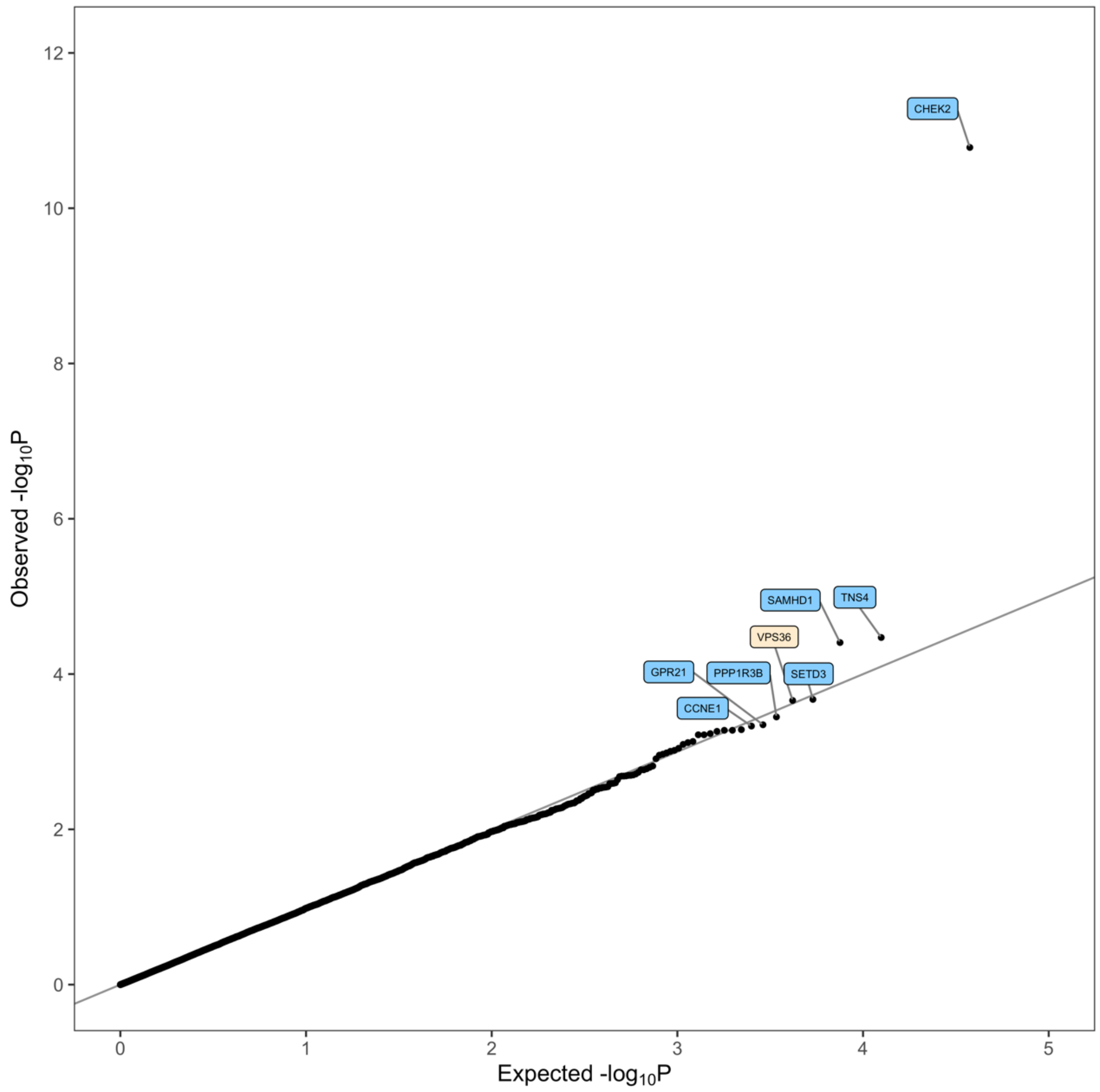


**Supplementary Figure 2 | Quantile-Quantile Plot of P-values from the meta-analysis assessing the association between rare missense variant carriers within genes and breast cancer risk.** Highlighted genes are those with $p<0.0005$. Blue corresponds to an increased risk of breast cancer, and cream corresponds to a decreased risk of breast cancer.

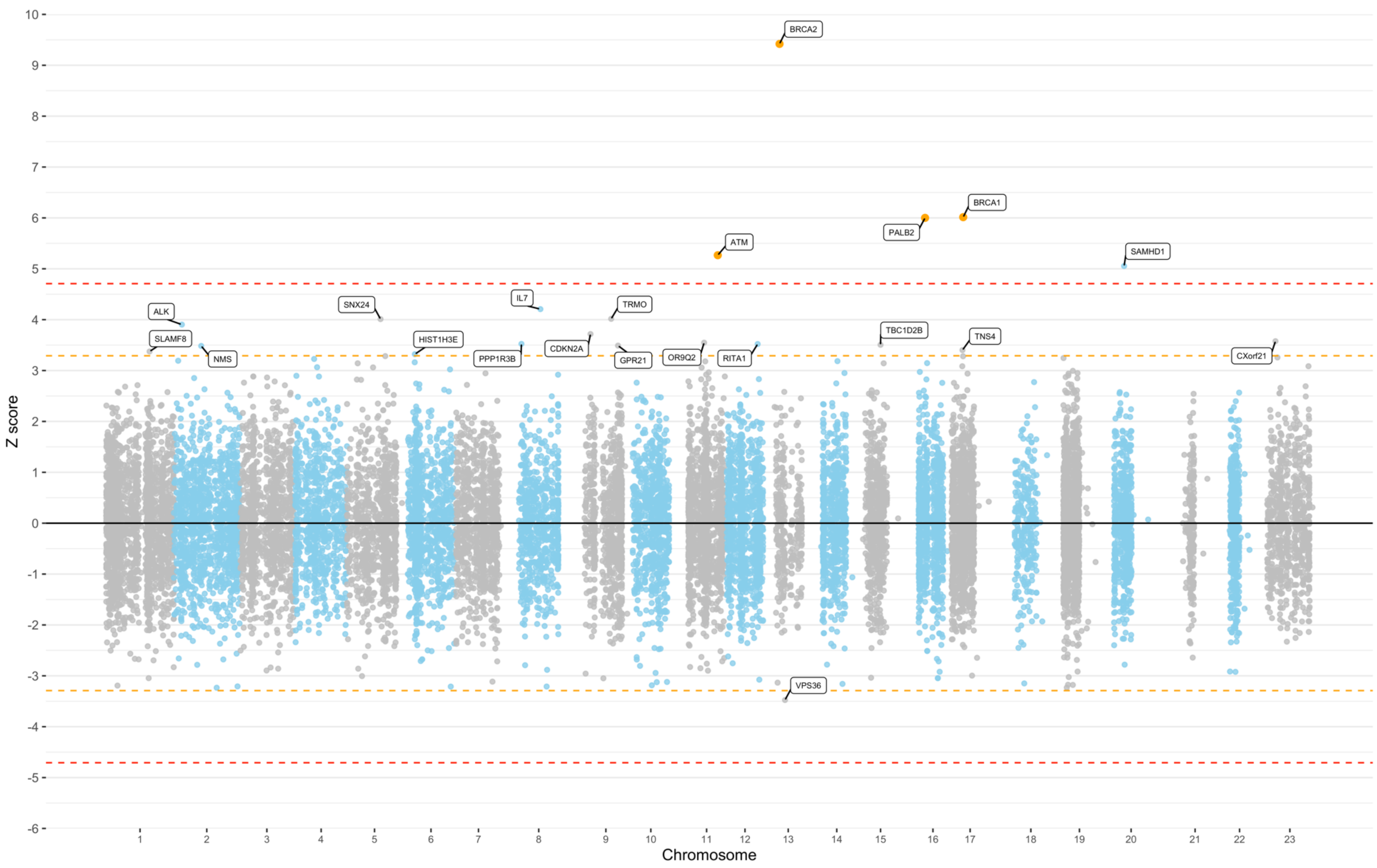


**Supplementary Figure 3 | Manhattan Plot of Z-scores from the meta-analysis assessing the association between PTV or deleterious rare missense variant carriers within genes and breast cancer risk.** Orange line corresponds to Z=±3.29, p=0.001 Red line corresponds to Z=±4.71, p=2.5x10-6

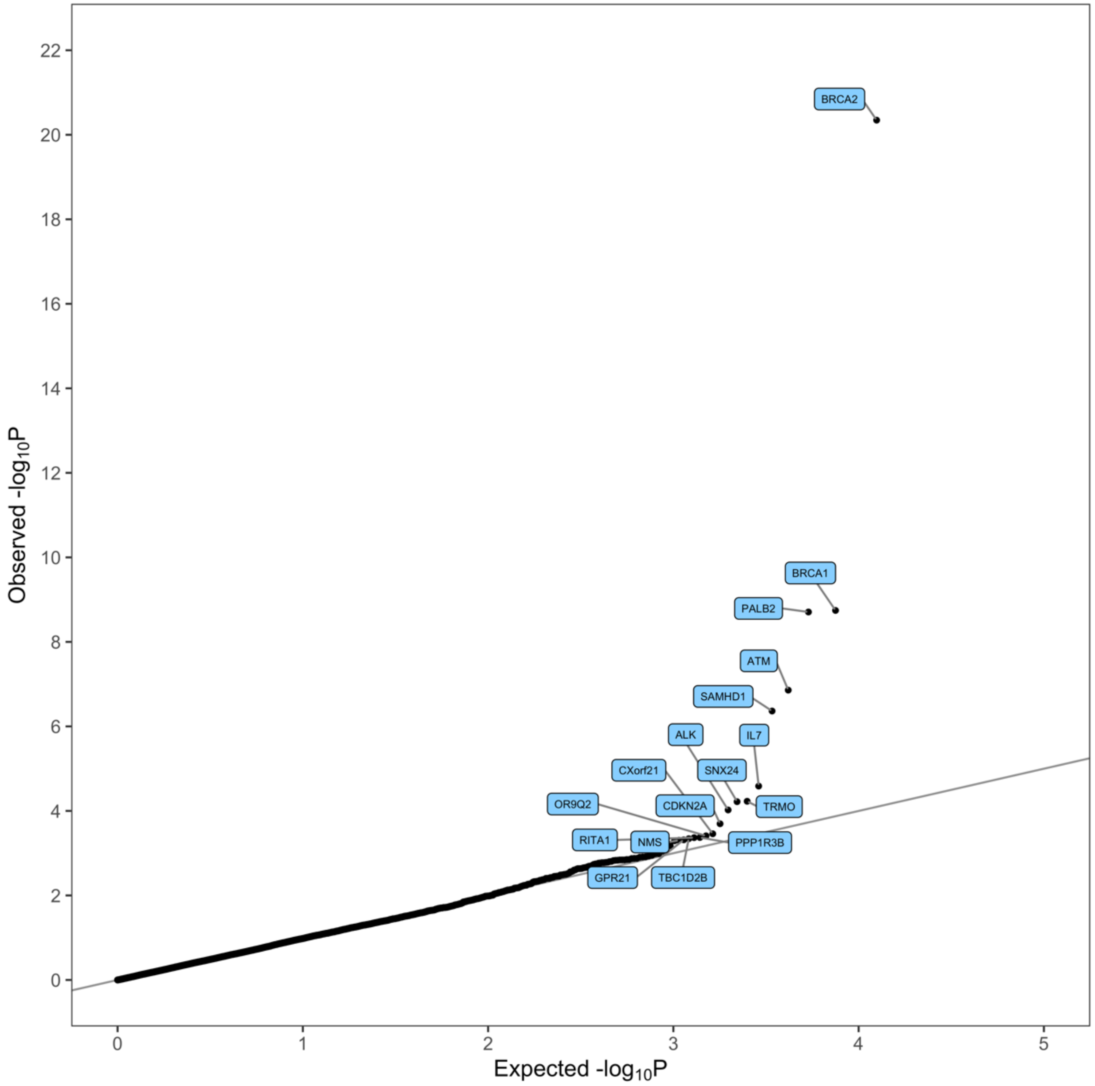


**Supplementary Figure 4 | Quantile-Quantile Plot of P-values from the Meta-Analysis assessing the association between PTV or deleterious rare missense variant carriers within genes and breast cancer risk.** All highlighted genes with $p<0.0005$ correspond to an increased risk of breast cancer.
