## Supplementary Methods for "Exome sequencing identifies novel susceptibility genes and defines the contribution of coding variants to breast cancer risk"

**Contribution of PTVs to the Familial Relative Risk**

As described in the main methods we assume a prior distribution for effect sizes (log-odds ratio) in which a proportion, $\alpha$, of genes are risk associated. For genes that are risk associated, the prior distribution for the log-relative risk is assumed to follow a negative exponential distribution. Thus, the log-relative risk $\beta$ has a density of the form $f(\beta|\alpha,\eta)$:

$\beta \sim \begin{cases} 0\ w.p.\ 1-\alpha \\ g(\beta|\eta)\ w.p.\ \alpha \end{cases}$, where $g(\beta|\eta) \sim \eta \exp(-\eta\beta)$

We derive an approximate likelihood for observed carrier counts separately for the BCAC dataset and UK biobank dataset, before combining them into one likelihood. This is maximised to estimate $\alpha$, $\beta$ and hence the posterior effect size distributions.

For the BCAC datasets, we consider a 2x2 contingency table of counts for each gene *j*:

| | **Control** | **Case** | **Total** |
|---|---|---|---|
| **Non-carrier** | $N_{B0} - n_{B0j}$ | $N_{B1} - n_{B1j}$ | $N_{B0} + N_{B1} - n_{Bj}$ |
| **Carrier** | $n_{B0j}$ | $n_{B1j}$ | $n_{B0j} + n_{B1j} = n_{Bj}$ |
| | $N_{B0}$ | $N_{B1}$ | $N_{B0} + N_{B1}$ |

Given the relative risk $e^{\beta_j}$ for gene j, and making the simplifying assumption that the frequency of pathogenic variants is low, the expected proportion of carriers that are cases is, to a good approximation, $P(Case\ |Carrier) = \frac{N_{B1}e^{\beta_j}}{N_{B0}+N_{B1}e^{\beta_j}}$.

Therefore, the number of case carriers, given the total number of carriers, can be modelled by a binomial distribution $n_{B1j} \sim Bin(n_{Bj}, \frac{N_{B1}e^{\beta_j}}{N_{B0}+N_{B1}e^{\beta_j}})$. Hence

$P(n_{B1j}|\ n_{Bj}, \beta_j) = \binom{n_{Bj}}{n_{B1j}} \left(\frac{N_{B1}e^{\beta_j}}{N_{B0}+N_{B1}e^{\beta_j}}\right)^{n_{B1j}} \left(1 - \frac{N_{B1}e^{\beta_j}}{N_{B0}+N_{B1}e^{\beta_j}}\right)^{n_{B0j}}$. Defining $\gamma_B = log(\frac{N_{B1}}{N_{B0}})$, this simplifies to

$$P\left(n_{B1j} \middle| n_{Bj}, \beta_j\right) = \binom{n_{Bj}}{n_{B1j}} \frac{\left(e^{\gamma_B}+1\right)^{n_{B0j}+n_{B1j}} e^{\beta_j n_{B1j}}}{\left(e^{\beta_j+\gamma_B}+1\right)^{n_{B0j}+n_{B1j}}}$$

For the UK Biobank dataset, we stratify the carrier counts by sex and family history status:

| | **FEMALE** | | | | | **MALE** | | | | |
|---|---|---|---|---|---|---|---|---|---|---|
| | Control | | Case | | | Control | | Case | | |
| | FH 0 | FH 1 | FH 0 | FH 1 | | FH 0 | FH 1 | FH 0 | FH 1 | |
| **Non-carrier** | $N_{F0} - n_{F0j}$ | $N_{F1} - n_{F1j}$ | $N_{F2} - n_{F2j}$ | $N_{F3} - n_{F3j}$ | $N_F - n_F$ | $N_{M0} - n_{M0j}$ | $N_{M1} - n_{M1j}$ | $N_{M2}$- $- n_{M2j}$ | $N_{M3} - n_{M3j}$ | $N_M - n_M$ |
| **Carrier** | $n_{F0j}$ | $n_{F1j}$ | $n_{F2j}$ | $n_{F3j}$ | $n_{F0j} + n_{F1j} + n_{F2j} + n_{F3j} = n_F$ | $n_{M0j}$ | $n_{M1j}$ | $n_{M2j}$ | $n_{M3j}$ | $n_{M0j} + n_{M1j} + n_{M2j} + n_{M3j} = n_M$ |
| | $N_{F0}$ | $N_{F1}$ | $N_{F2}$ | $N_{F3}$ | $N_{F0} + N_{F1} + N_{F2} + N_{F3} = N_F$ | $N_{M0}$ | $N_{M1}$ | $N_{M2}$ | $N_{M3}$ | $N_{M0} + N_{M1} + N_{M2} + N_{M3} = N_M$ |

Thus, for each sex (subscript F or M), the phenotype of an individual has four possibilities (control +/- family history, case +/- family history). The probabilities of these phenotypes for a carrier are given by:

$$P(phenotype = k|carrier) = \frac{N_k e^{\beta_k j}}{N_0 + N_1 e^{\beta_1 j} + N_2 e^{\beta_2 j} + N_3 e^{\beta_3 j}}$$

Where $e^{\beta_k j}$ is the relative risk of phenotype *k*, relative to phenotype 0. For a rare dominant disease allele, and assuming that positive family history is relatively rare, $\boldsymbol{e^{\beta_{1j}} \approx \frac{1}{2}(e^{\beta_j} + 1)}$, $\boldsymbol{e^{\beta_{2j}} \approx e^{\beta_j}}$, $\boldsymbol{e^{\beta_{3j}} \approx \frac{1}{2}(3e^{\beta_j} - 1)}$. That is, $e^{\beta_{kj}} \approx \frac{1}{2}\left(ke^{\beta_j} + 2 - k\right)$ [1]

Therefore, for each sex, the number of carriers in each stratum can be modelled by a multinomial distribution with a probability mass function:

$$n_{oj}, n_{1j}, n_{2j}, n_{3j} \sim Multinom\left(n_j, \frac{N_0}{N_0+\frac{1}{2}N_1 \mathrm{e}^{\beta_j}+\frac{1}{2}N_1+N_2 e^{\beta_j}+\frac{3}{2}N_3 \mathrm{e}^{\beta_j}-\frac{1}{2}N_3}, \frac{\frac{1}{2}N_1 \mathrm{e}^{\beta_j}+\frac{1}{2}N_1}{N_0+\frac{1}{2}N_1 \mathrm{e}^{\beta_j}+\frac{1}{2}N_1+N_2 e^{\beta_j}+\frac{3}{2}N_3 \mathrm{e}^{\beta_j}-\frac{1}{2}N_3}, \frac{N_2 e^{\beta_j}}{N_0+\frac{1}{2}N_1 \mathrm{e}^{\beta_j}+\frac{1}{2}N_1+N_2 e^{\beta_j}+\frac{3}{2}N_3 \mathrm{e}^{\beta_j}-\frac{1}{2}N_3}, \frac{\frac{3}{2}N_3 \mathrm{e}^{\beta_j}-\frac{1}{2}N_3}{N_0+\frac{1}{2}N_1 \mathrm{e}^{\beta_j}+\frac{1}{2}N_1+N_2 e^{\beta_j}+\frac{3}{2}N_3 \mathrm{e}^{\beta_j}-\frac{1}{2}N_3}\right)$$

.

So that:

$$P\left(n_{oj}, n_{1j}, n_{2j}, n_{3j} | n_j, \beta_j\right) = \frac{\prod_{k=0}^{3}\left(N_k \frac{1}{2}\left(ke^{\beta_j}+2-k\right)\right)^{n_{kj}}}{\left(\sum_{k=0}^{3} N_k \frac{1}{2}\left(ke^{\beta_j}+2-k\right)\right)^{n_j}}$$

Defining $\gamma_{Fk} = log(\frac{N_{Fk}}{N_{F0}}), \gamma_{Mk} = log(\frac{N_{Mk}}{N_{M0}})$, and multiplying the probabilities for males and females, this simplifies to:

$$P\left(n_{oj}, n_{1j}, n_{2j}, n_{3j} | n_j, \beta_j\right) = C \frac{\prod_{k=0}^{3}\left(\frac{1}{2}\left(ke^{\beta_j}+2-k\right)\right)^{n_{Fkj}} \prod_{k=0}^{3}\left(\frac{1}{2}\left(ke^{\beta_j}+2-k\right)\right)^{n_{Mkj}}}{\left(\sum_{k=0}^{3} e^{\gamma_{F_k}} \frac{1}{2}\left(ke^{\beta_j}+2-k\right)\right)^{n_{Fj}} \left(\sum_{k=0}^{3} e^{\gamma_{M_k}} \frac{1}{2}\left(ke^{\beta_j}+2-k\right)\right)^{n_{Mj}}}$$

Where $C = \binom{n_{Fj}}{n_{F0j} \quad n_{F1j} \quad n_{F2j} \quad n_{F3j}} \binom{n_{Mj}}{n_{M0j} \quad n_{M1j} \quad n_{M2j} \quad n_{M3j}} e^{\sum_{k=0}^{3}\gamma_{Fk}n_{Fkj}+\sum_{k=0}^{3}\gamma_{Mk}n_{Mkj}}$ is independent of the prior distribution.

The likelihoods for the BCAC dataset and the UK biobank dataset are then multiplied and integrated over the prior distribution to give the likelihood to be maximised:

$$L(\alpha,\eta) \propto \prod_{j=1}^{J} \int \frac{e^{\beta_j n_{B1j}}}{(e^{\beta_j+\gamma_B}+1)^{n_{B0j}+n_{B1j}}} \frac{\prod_{k=0}^{3}\left(\frac{1}{2}\left(ke^{\beta_j}+2-k\right)\right)^{n_{Fkj}} \prod_{k=0}^{3}\left(\frac{1}{2}\left(ke^{\beta_j}+2-k\right)\right)^{n_{Mkj}}}{\left(\sum_{k=0}^{3} e^{\gamma_{F_k}} \frac{1}{2}\left(ke^{\beta_j}+2-k\right)\right)^{n_{Fj}} \left(\sum_{k=0}^{3} e^{\gamma_{M_k}} \frac{1}{2}\left(ke^{\beta_j}+2-k\right)\right)^{n_{Mj}}} f\left(\beta_j|\alpha,\eta\right) d\beta_j$$

Where $f(\beta_j|\alpha,\eta)$ is the prior distribution on $\beta_j$.

Writing: $L_j(\beta_j) = (e^{\gamma_B}+1)^{n_{B0j}+n_{B1j}} \left(\sum_{k=0}^{3} e^{\gamma_{F_k}}\right)^{n_{Fj}} \left(\sum_{k=0}^{3} e^{\gamma_{M_k}}\right)^{n_{Mj}} \frac{e^{\beta_j n_{B1j}}}{\left(e^{\beta_j+\gamma_B}+1\right)^{n_{B0j}+n_{B1j}}} \frac{\prod_{k=0}^{3}\left(\frac{1}{2}\left(ke^{\beta_j}+2-k\right)\right)^{n_{Fkj}} \prod_{k=0}^{3}\left(\frac{1}{2}(ke^{\beta_j}+2-k)\right)^{n_{Mkj}}}{\left(\sum_{k=0}^{3} e^{\gamma_{F_k}} \frac{1}{2}\left(\left(ke^{\beta_j}+2-k\right)\right)\right)^{n_{Fj}} \left(\sum_{k=0}^{3} e^{\gamma_{M_k}} \frac{1}{2}(ke^{\beta_j}+2-k)\right)^{n_{Mj}}}$

$L(\alpha,\eta) \propto \prod_{j=1}^{J} \int L_j\left(\beta_j\right) f\left(\beta_j|\alpha,\eta\right) d\beta_j = \prod_{j=1}^{J}\left(1-\alpha+\alpha \int L_j\left(\beta_j\right) g\left(\beta_j|\eta\right) d\beta_j\right) = \prod_{j=1}^{J}\left(1-\alpha+\alpha L_{*j}\right)$ say.

A major advantage of this approach, which conditions on the total carrier count, is that it avoids the problems of estimating the allele frequency for each gene simultaneously with the relative risk parameters so that only maximisation over $\alpha$and $\eta$ is required. This is relatively straightforward.

The posterior probability a gene is associated, given the estimates of $\alpha$and $\eta$, is then:

$$P(\beta_j|Data) = \frac{\alpha \int L_j(\beta_j) g(\beta_j|\eta) d\beta_j}{1 - \alpha + \alpha \int L_j(\beta_j) g(\beta_j|\eta) d\beta_j} = \frac{\alpha L_{*j}}{1 - \alpha + \alpha L_{*j}}$$

The posterior mean $\beta_j$ is given by:

$$\frac{\alpha \int \beta_j L_j(\beta_j) g(\beta_j|\eta) d\beta_j}{1 - \alpha + \alpha L_{*j}}$$

And the posterior mean relative risk $e^{\beta_j}$ is given by:

$$\frac{\alpha \int e^{\beta_j} L_j(\beta_j) g(\beta_j|\eta) d\beta_j}{1 - \alpha + \alpha L_{*j}}$$

To calculate the <u>contribution of PTVs to the FRR</u>, we note that for genes with aggregate PTV frequency, $p_j$, associated with relative risk $e^{\beta_j}$, the FRR is:

$$\lambda_j = 1 + \frac{p_j(e^{\beta_j} - 1)^2}{(2p_j(e^{\beta_j} - 1) + 1)^2}$$

This requires an estimate of the allele frequency for each gene. The simplest approach is to use the carrier frequencies in controls, thus:

$$\hat{p}_{Aj} = \frac{(n_{F0j} + n_{F1j} + n_{B0j})}{2(N_{F0} + N_{F1} + N_{B0})}$$

Hence:

$$\lambda_{jA} = 1 + \frac{\alpha}{1 - \alpha + \alpha L_{*j}} \int L_j(\beta_j) g(\beta_j|\eta) \frac{p_{jA}(e^{\beta_j} - 1)^2}{(2p_{jA}(e^{\beta_j} - 1) + 1)^2} d\beta_j$$

A potentially better approach is to also utilise the case data, estimating the allele frequency based on the posterior distribution of the relative risk. Thus:

$$p_{Bj}(\beta_j) = \frac{n_{F0j} + n_{F1j} + n_{F2j} + n_{F3j} + n_{B0j} + n_{B1j}}{2(N_{F0} + N_{F1} + N_{B0} + e^{\beta_j}(N_{F2} + N_{F3} + N_{B1})}$$

Hence:

$$\lambda_{jB} = 1 + \frac{\alpha}{1 - \alpha + \alpha L_{*j}} \int L_j(\beta_j) g(\beta_j|\eta) \frac{p_{jB}(\beta_j)(e^{p_{jB}(\beta_j)} - 1)^2}{(2p_{jB}(\beta_j)(e^{p_{jB}(\beta_j)} - 1) + 1)^2} d\beta_j$$

In the main analyses, we used the second method. However, for *BRCA1, BRCA2* and *PALB2* replaced the allele frequency estimates with external estimates from the literature, namely the estimates used in the current default BOADICEA/Canrisk model[2]. This is to account for the fact that the allele frequency estimates in the dataset will be underestimated (due to the deliberate exclusion of known carriers in the case of the BCAC exome sequencing, and potential under-ascertainment of carriers in UK Biobank).

The total FRR over all genes, assuming an additive model, is then given by:

$$\hat{\lambda}_{TOT} = 1 + \sum_{j=1}^{J} (\lambda_j - 1)$$

Assuming that the PTVs combined multiplicatively with other genetic or familial factors, and an overall FRR of 2, the percentage contribution of each gene to the overall FRR is therefore: $100 \times \frac{log(\hat{\lambda}_j)}{log(2)}$ and the total contribution of PTVs in all genes is: $100 \times \frac{log(\hat{\lambda}_{TOT})}{log(2)}$.


1. Risch, N. Linkage strategies for genetically complex traits. I. Multilocus models. *American journal of human genetics* **46**, 222-228 (1990).
2. Lee, A. *et al.* Enhancing the BOADICEA cancer risk prediction model to incorporate new data on RAD51C, RAD51D, BARD1, updates to tumour pathology and cancer incidences. (Cold Spring Harbor Laboratory, 2022).
