## Supplementary Note for "Exome sequencing identifies novel susceptibility genes and defines the contribution of coding variants to breast cancer risk"

**Study-specific Funding**

The **ABCS** study was supported by the Dutch Cancer Society [grants NKI 2007-3839; 2009 4363]. The BREast Oncology GAlician Network (**BREOGAN**) is funded by Acción Estratégica de Salud del Instituto de Salud Carlos III FIS PI12/02125/Cofinanciado and FEDER PI17/00918/Cofinanciado FEDER; Acción Estratégica de Salud del Instituto de Salud Carlos III FIS Intrasalud (PI13/01136); Programa Grupos Emergentes, Cancer Genetics Unit, Instituto de Investigacion Biomedica Galicia Sur. Xerencia de Xestion Integrada de Vigo-SERGAS, Instituto de Salud Carlos III, Spain; Grant 10CSA012E, Consellería de Industria Programa Sectorial de Investigación Aplicada, PEME I + D e I + D Suma del Plan Gallego de Investigación, Desarrollo e Innovación Tecnológica de la Consellería de Industria de la Xunta de Galicia, Spain; Grant EC11-192. Fomento de la Investigación Clínica Independiente, Ministerio de Sanidad, Servicios Sociales e Igualdad, Spain; and Grant FEDER-Innterconecta. Ministerio de Economia y Competitividad, Xunta de Galicia, Spain. The **CECILE** study was supported by Fondation de France, Institut National du Cancer (INCa), Ligue Nationale contre le Cancer, Agence Nationale de Sécurité Sanitaire, de l'Alimentation, de l'Environnement et du Travail (ANSES), Agence Nationale de la Recherche (ANR). The **GC-HBOC** (German Consortium of Hereditary Breast and Ovarian Cancer) is supported by the German Cancer Aid (grant no 110837 and 70114178, coordinator: Rita K. Schmutzler, Cologne) and the Federal Ministry of Education and Research, Germany (grant no 01GY1901). This work was also funded by the European Regional Development Fund and Free State of Saxony, Germany (LIFE - Leipzig Research Centre for Civilization Diseases, project numbers 713-241202, 713-241202, 14505/2470, 14575/2470). The **GESBC** was supported by the Deutsche Krebshilfe e. V. [70492] and the German Cancer Research Center (DKFZ). The **HABCS** study was supported by the Claudia von Schilling Foundation for Breast Cancer Research, by the Lower Saxonian Cancer Society, and by the Rudolf Bartling Foundation. The **MARIE** study was supported by the Deutsche Krebshilfe e.V. [70-2892-BR I, 106332, 108253, 108419, 110826, 110828], the Hamburg Cancer Society, the German Cancer Research Center (DKFZ) and the Federal Ministry of Education and Research (BMBF) Germany [01KH0402]. The **MASTOS** study was supported by “Cyprus Research Promotion Foundation” grants 0104/13 and 0104/17, and the Cyprus Institute of Neurology and Genetics. **MYBRCA** is funded by research grants from the Wellcome Trust (v203477/Z/16/Z), the Malaysian Ministry of Higher Education (UM.C/HlR/MOHE/06) and Cancer Research Malaysia. **SEARCH** is funded by Cancer Research UK [C490/A10124, C490/A16561] and supported by the UK National Institute for Health Research Biomedical Research Centre at the University of Cambridge. The University of Cambridge has received salary support for PDPP from the NHS in the East of England through the Clinical Academic Reserve. **SGBCC** is funded by the National Research Foundation Singapore, NUS start-up Grant, National University Cancer Institute Singapore (NCIS) Centre Grant, Breast Cancer Prevention Programme, Asian Breast Cancer Research Fund and the NMRC Clinician Scientist Award (SI Category). Population-based controls were from the Multi-Ethnic Cohort (MEC) funded by grants from the Ministry of Health, Singapore, National University of Singapore and National University Health System, Singapore.

**Acknowledgments**

**ABCS** thanks the Blood bank Sanquin, The Netherlands. The **BREOGAN** study would not have been possible without the contributions of the following: Manuela Gago-Dominguez, Jose Esteban Castelao, Angel Carracedo, Victor Muñoz Garzón, Alejandro Novo Domínguez, Maria Elena Martinez, Sara Miranda Ponte, Carmen Redondo Marey, Maite Peña Fernández, Manuel Enguix Castelo, Maria Torres, Manuel Calaza (BREOGAN), José Antúnez, Máximo Fraga and the staff of the Department of Pathology and Biobank of the University Hospital Complex of Santiago-CHUS, Instituto de Investigación Sanitaria de Santiago, IDIS, Xerencia de Xestion Integrada de Santiago-SERGAS; Joaquín

González-Carreró and the staff of the Department of Pathology and Biobank of University Hospital Complex of Vigo, Instituto de Investigacion Biomedica Galicia Sur, SERGAS, Vigo, Spain. **HABCS** thanks Michael Bremer. **MARIE** thanks Petra Seibold, Nadia Obi, Sabine Behrens, Ursula Eilber and Muhabbet Celik. **MASTOS** thanks all the study participants and express appreciation to the doctors: Yiola Marcou, Eleni Kakouri, Panayiotis Papadopoulos, Simon Malas and Maria Daniel, as well as to all the nurses and volunteers who provided valuable help towards the recruitment of the study participants. **MYBRCA** thanks study participants and research staff (particularly Patsy Ng, Nurhidayu Hassan, Yoon Sook-Yee, Daphne Lee, Lee Sheau Yee, Phuah Sze Yee and Norhashimah Hassan) for their contributions and commitment to this study. We thank the **SEARCH** and EPIC teams. **SGBCC** thanks the participants and all research coordinators for their excellent help with recruitment, data and sample collection.
